# Observational study of changes in utilization and outcomes in non-invasive ventilation in COVID-19

**DOI:** 10.1101/2021.03.28.21254477

**Authors:** Christian Karagiannidis, Corinna Hentschker, Michael Westhoff, Steffen Weber-Carstens, Uwe Janssens, Stefan Kluge, Michael Pfeifer, Claudia Spies, Tobias Welte, Rolf Rossaint, Carina Mostert, Wolfram Windisch

## Abstract

**Rationale:** The role of non-invasive ventilation (NIV) in severe COVID-19 remains a matter of debate.

**Objectives:** To determine the utilization and outcome of NIV in COVID-19 in an unbiased cohort.

**Methods:** Observational study of confirmed COVID-19 cases of claims data of the Local Health Care Funds comparing patients with non-invasive and invasive mechanical ventilation (IMV) between spring versus autumn period 2020.

**Measurements and Main Results:** Nationwide cohort of 7490 cases (median/IQR age 70/60–79 years, 66% male) 3851 (51%) patients primarily received IMV without NIV, 1614 (22%) patients received NIV without subsequent intubation, and 1247 (17%) patients had NIV failure (NIV-F), defined by subsequent endotracheal intubation. The proportion of patients who received invasive MV decreased from 74% to 39% during the second period. Accordingly, the proportion of patients with NIV exclusively increased from 10% to 28%, and those failing NIV increased from 9% to 21%. Median length of hospital stay decreased from 26 to 22 days, and duration of MV decreased from 11.6 to 7.6 days. The NIV failure rate decreased from 49% to 42%. Overall mortality remained unchanged (51% versus 53%). Mortality was 39% with NIV-only, 52% with IMV and 66% with NIV-F with mortality rates steadily increasing from 58% in early NIV-F (day 1) to 75% in late NIV-F (>4 days).

**Conclusion:** Utilization of NIV rapidly increased during the autumn period, which was associated with a reduced duration of MV, but not with overall mortality. High NIV-F rates are associated with increased mortality, particularly in late NIV-F.

**Funding:** Institutional support and physical resources were provided by the University Witten/Herdecke and Kliniken der Stadt Köln and the Federal Association of the Local Health Care Funds.

**At a Glance Commentary:** *Scientific Knowledge on the Subject:* Current management of ventilatory support in COVID-19 patients with respiratory failure is heterogeneous. Despite increasing use of non-invasive ventilation (NIV), defining intubation criteria still remains a matter of uncertainty and discussion, especially with regard to the balance between the NIV benefits and the risk of NIV failure. In addition, robust data concerning the influence of the duration and failure of NIV on intubation and mortality rates are still missing, although the time span between initiation of NIV and subsequent intubation in case of respiratory failure progression is suggested to influence patient outcome.

*What This Study Adds to the Field:* This is the first large observational study describing differences of ventilatory strategies between the spring and autumn period of the SARS-CoV-2 pandemic in Germany and provides the in-hospital mortality rate of 7,490 patients who received mechanical ventilation. The increased utilization of NIV from 10% (first period) to 29% (second period) was associated with overall reduced durations of mechanical ventilation and length of hospital stay, but overall mortality remained comparably high and reached 51%, 53% respectively. Patients succeeding with NIV had lower mortality rates than those getting intubated without preceding NIV attempts, but those failing NIV had higher mortality rates, respectively, and this became even more predominant in late NIV failure. The present observational study shows the increasing role of NIV in the concert of ICU medicine related to COVID-19, but also clearly addresses its risks in addition to its benefits, both impacting on mortality.

## Introduction

Within one year, the SARS-CoV-2 pandemic has affected more than 125 million people worldwide. Mortality rates of patients requiring ICU treatment are ranging up to over 50% (1-4), depending on the severity of respiratory failure and response to treatment, but also influenced by age, comorbidities and a ceiling of therapeutic interventions (1, 2, 5-7).

Mechanical ventilation (MV) is a life-saving option in severe COVID-19 cases, but mortality rates in patients on MV remain high (4, 5). Non-invasive ventilation (NIV) is suggested to reduce the complications of invasive MV(8). For COVID-19 patients, current guidelines recommend stepping up to NIV when oxygenation worsens during oxygen therapy, and to consider intubation if PaO_2_/FiO_2_ is decreased below 150 mmHg (9-11) or the clinical presentation of the patients has worsened (9, 11-15). However, global current practices of MV widely differ, also depending on COVID-19-associated limited resources (4, 16, 17). Therefore, the role of NIV remains a matter of uncertainty and discussion, especially with regard to the balance between the NIV benefits and the risk of NIV failure (NIV-F). The mortality of patients receiving NIV was in a wide range up to 45% (1, 18). In contrast to this, the mortality rate in patients with NIV-F ranged between 35% and 74% (18-20). Hence, interpretation of data and obtaining conclusive strategies concerning the optimal and individual timing of intubation remain uncertain (21).

Therefore, the aim of the current study was to determine detailed characteristics and outcomes of 7,490 hospitalized COVID-19 patients with MV in a large, unselected and unbiased cohort of patients with confirmed COVID-19 in one of the least resource limited health care systems, particularly focusing on patients requiring invasive MV or NIV with specific emphasis on NIV-F.

## Data and methods

The inpatient data of the general local health insurance funds, which cover around a third of the German population, were analyzed. It is an administrative data set containing patient information like age, gender, diagnosis and procedure codes. However, detailed medical information such as laboratory finding is not recorded. All cases were included for which admission and discharge dates as well as diagnoses and procedures were coded. Only patients with laboratory-confirmed SARS-CoV-2 infection (diagnosis code U07.1!) were included. The patients were at least 18 years old and were admitted to hospital between February 1, 2020 and November 30, 2020.

The original data structure is at the case level, i.e. insured persons who were transferred to another hospital during their hospital stay appear several times in the data set. Therefore, cases who were transferred during their hospital stay (discharge date of one hospital corresponds to the admission date of another hospital) were merged. Thus, the current analysis was performed at the patient level.

The primary analysis includes all patients with mechanical ventilation, but secondary analysis focuses on patients with MV for more than 6 hours, i.e. invasive MV or NIV. These patients were divided into three subgroups: 1) patients with primary invasive MV without any NIV attempt preceding intubation, 2) patients with NIV exclusively, who have not been escalated to intubation, and 3) those with NIV-F, defined by endotracheal intubation following NIV. In the last group, a procedure code for both NIV and invasive MV was assigned. If invasive MV was started at least one day later than NIV, the patient was assigned to the NIV-F group. If invasive ventilation was started on the same day as NIV initiation, the patient was assigned to the invasive MV. Both, patients with less than 6 documented hours of ventilation and patients with more than 6 documented hours of ventilation but without a corresponding procedure code for NIV or invasive ventilation were not assigned to the three subgroups. With the inclusion of the procedural data, it was possible to roughly determine the NIV duration when switching from NIV to invasive MV.

Due to the timeliness of the data, some patients are missing in the more recent months (admission in October and November). These patients have been hospitalized for a long time and no claim data are yet available. Based on the experience from the first period of the pandemic, these are patients with lower mortality rates and a longer duration of MV, but this group consists of only a few patients. The study was approved by the local ethical committee (University Witten/Herdecke, 92/2020).

## Findings

Between February 1 and November 30, 7,490 hospitalized Covid-19 patients received MV, 32% during the first wave of the pandemic (February to May) and 59% during the second wave (October to November) (Table1). Data of patients treated during the summer months (June to September) is not shown separately in the table due to the relatively low number (N=643). Age distribution, sex and the frequency of comorbidities show only slight differences when comparing the two periods of the pandemic as shown in Table 1. The overall median length of hospital stay has decreased from 26 days during the first wave of the pandemic to 22 days during the second wave. This also applies to the overall duration of MV, which decreased from 11.6 to 7.6 days, respectively.

**Table 1.** Patient characteristics comparing the spring and autumn period, ECMO=extracorporeal membrane oxygenation,

|  | Patients by month of hospital admission |  |  |
| --- | --- | --- | --- |
|  | Total | Admission between February and May | Admission between October and November |
| Number of patients | 7490 | 2419 | 4428 |
| Age (years) |  |  |  |
| Mean (SD) | 68.5 (13.2) | 68.1 (13.4) | 69.2 (13.0) |
| Median (IQR) | 70.0 (60.0, 79.0) | 70.0 (60.0, 79.0) | 71.0 (61.0, 79.0) |
| 18-49 years | 625 (8.3%) | 208 (8.6%) | 328 (7.4%) |
| 50-59 years | 1,115 (14.9%) | 386 (16.0%) | 622 (14.0%) |
| 60-69 years | 1,812 (24.2%) | 564 (23.3%) | 1,075 (24.3%) |
| 70-79 year | 2,217 (29.6%) | 741 (30.6%) | 1,314 (29.7%) |
| ≥ 80 years | 1,721 (23.0%) | 520 (21.5%) | 1,089 (24.6%) |
| Male | 4,913 (65.6%) | 1,599 (66.1%) | 2,871 (64.8%) |
| Female | 2,577 (34.4%) | 820 (33.9%) | 1,557 (35.2%) |
| Elixhauser comorbidities |  |  |  |
| Hypertension | 5,032 (67.2%) | 1,567 (64.8%) | 3,029 (68.4%) |
| Diabetes | 3,181 (42.5%) | 969 (40.1%) | 1,951 (44.1%) |
| Cardiac arrhythmias | 3,188 (42.6%) | 1,103 (45.6%) | 1,826 (41.2%) |
| Renal failure | 2,045 (27.3%) | 638 (26.4%) | 1,260 (28.5%) |
| Congestive heart failure | 2,527 (33.7%) | 817 (33.8%) | 1,480 (33.4%) |
| Chronic pulmonary disease | 1,477 (19.7%) | 485 (20.0%) | 864 (19.5%) |
| Obesity | 1,128 (15.1%) | 359 (14.8%) | 665 (15.0%) |
| Patients transferred between hospitals | 2,554 (34.1%) | 868 (35.9%) | 1,390 (31.4%) |
| Length of hospital stay (days) |  |  |  |
| Mean (SD) | 33.4 (30.9) | 36.3 (35.8) | 29.5 (23.8) |
| Median (IQR) | 24.0 (13.0, 43.0) | 26.0 (13.0, 49.0) | 22.0 (13.0, 39.0) |
| Ventilation (days) |  |  |  |
| Mean (SD) | 14.2 (16.3) | 17.1 (18.6) | 12.2 (13.8) |
| Median (IQR) | 9.0 (2.7, 19.6) | 11.6 (4.7, 23.2) | 7.6 (2.0, 17.3) |
| Tracheostomy | 1,894 (25.3%) | 745 (30.8%) | 956 (21.6%) |
| ECMO | 571 (7.6%) | 189 (7.8%) | 306 (6.9%) |
| Dialysis | 1,801 (24.0%) | 726 (30.0%) | 909 (20.5%) |
| Type of ventilation |  |  |  |
| Invasive ventilation only (IMV) | 3,851 (51.4%) | 1,791 (74.0%) | 1,719 (38.8%) |
| Non-invasive ventilation only (NIV) | 1,614 (21.5%) | 230 (9.5%) | 1,258 (28.4%) |
| Non-invasive ventilation failure (NIV-F) | 1,247 (16.6%) | 219 (9.1%) | 927 (20.9%) |
| Duration of ventilation between 1-6 hours | 473 (6.3%) | 92 (3.8%) | 335 (7.6%) |
| No ventilation procedure code | 305 (4.1%) | 87 (3.6%) | 189 (4.3%) |
| In-hospital mortality | 3,819 (51.0%) | 1,223 (50.6%) | 2,335 (52.7%) |

A major difference between the spring and autumn period of the pandemic refers to the application of the different MV modalities. During the first period, 74% of the patients received invasive MV directly without having previously received NIV as a first escalation step. In contrast, only 39% received immediate invasive MV during the second pandemic wave. Consequently, more patients were escalated from oxygen therapy to NIV during the second period (Table 1) with both patients successfully treated with NIV increasing from 10% to 28% and those with NIV-F increasing from 9% to 21%. However, the NIV-F rate decreased from 49% (219 of 449) to 42% (927 of 2,185).

The overall mortality rate of patients receiving any form of MV in the first and second wave of the pandemic remained stable with 51% and 53%. Overall mortality rates were lower for patients receiving NIV only (39%) compared to those with invasive MV only (52%), as illustrated in more detail in Figure 1A. However, mortality rates of patients with NIV-F were highest (66%). Of note, the mortality rate in patients with NIV-F increased steadily, from 58% in patients with NIV-F on the first day to 75% in those with NIV-F on day 5 or later (Figure 1B).

**Figure 1.**
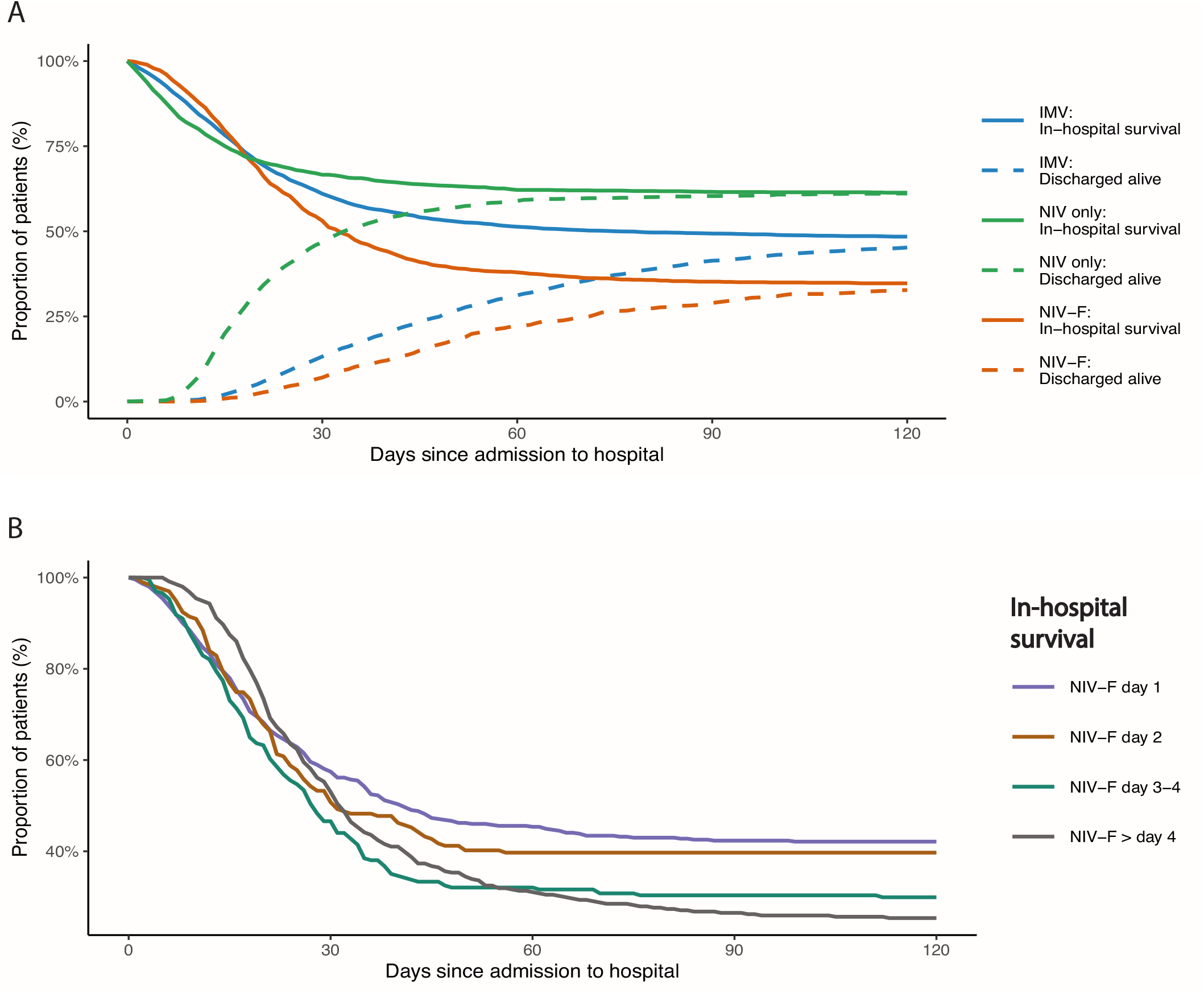
A. In-hospital mortality by type of ventilation. IMV=invasive mechanical ventilation (n=3851), NIV = non-invasive-ventilation failure (n=1614) and NIV-F = non-invasive-ventilation failure (n=1247). B. In-hospital mortality of non-invasive-ventilation failure (NIV-F, n=1247) by day of intubation

Overall, 2,861 patients had initially received NIV, with 1,247 having failed (NIV-F rate 44%) (Table 2). The highest proportion of NIV-F was found in the age group between 60–69 years (54%), while the age group between 70–79 years had the lowest failure rate (31%). The NIV-F rate was lower in women (37%) than in men (47%). There was no clear trend for the influence of comorbidities both on the decision to intubate the patient immediately and on the risk of NIV-F, i.e. the proportion of patients with a specific comorbidity was similar in both groups. One exception refers to cardiac arrhythmias, which were lowest in patients successfully treated with NIV (Table 2).

**Table 2.** Patient characteristics by type of ventilation. ECMO=extracorporeal membrane oxygenation, NIV=non-invasive ventilation, NIV-F= non-invasive ventilation failure.

|  | Invasive ventilation only | Non-invasive ventilation only | Non-invasive ventilation failure | Non-invasive ventilation failure by day of failure |  |  |  |
| --- | --- | --- | --- | --- | --- | --- | --- |
|  |  |  |  | First day | Second day | Third or fourth day | Fifth day or later |
| Number of patients | 3851 | 1614 | 1247 | 463 | 199 | 234 | 351 |
| Age (years) |  |  |  |  |  |  |  |
| Mean (SD) | 67.5 (13.1) | 70.2 (13.7) | 68.3 (11.9) | 67.3 (12.6) | 68.5 (13.1) | 68.4 (11.1) | 69.6 (10.8) |
| Median (IQR) | 69.0 (59.0, 78.0) | 73.0 (61.0, 81.0) | 70.0 (61.0, 78.0) | 69.0 (60.0, 78.0) | 71.0 (61.0, 79.0) | 70.0 (62.0, 76.8) | 72.0 (64.0, 78.0) |
| 18-49 years | 349 (9.1%) | 128 (7.9%) | 87 (7.0%) | 39 (8.4%) | 17 (8.5%) | 13 (5.6%) | 18 (5.1%) |
| 50-59 years | 620 (16.1%) | 224 (13.9%) | 157 (12.6%) | 70 (15.1%) | 21 (10.6%) | 29 (12.4%) | 37 (10.5%) |
| 60-69 years | 987 (25.6%) | 310 (19.2%) | 364 (29.2%) | 129 (27.9%) | 58 (29.1%) | 74 (31.6%) | 103 (29.3%) |
| 70-79 year | 1,166 (30.3%) | 441 (27.3%) | 409 (32.8%) | 144 (31.1%) | 57 (28.6%) | 76 (32.5%) | 132 (37.6%) |
| ≥ 80 years | 729 (18.9%) | 511 (31.7%) | 230 (18.4%) | 81 (17.5%) | 46 (23.1%) | 42 (17.9%) | 61 (17.4%) |
| Male | 2,564 (66.6%) | 992 (61.5%) | 877 (70.3%) | 320 (69.1%) | 147 (73.9%) | 161 (68.8%) | 249 (70.9%) |
| Female | 1,287 (33.4%) | 622 (38.5%) | 370 (29.7%) | 143 (30.9%) | 52 (26.1%) | 73 (31.2%) | 102 (29.1%) |
| Elixhauser comorbidities |  |  |  |  |  |  |  |
| Hypertension | 2,555 (66.3%) | 1,115 (69.1%) | 877 (70.3%) | 318 (68.7%) | 144 (72.4%) | 156 (66.7%) | 259 (73.8%) |
| Diabetes | 1,653 (42.9%) | 688 (42.6%) | 558 (44.7%) | 210 (45.4%) | 88 (44.2%) | 106 (45.3%) | 154 (43.9%) |
| Cardiac arrhythmias | 1,718 (44.6%) | 566 (35.1%) | 597 (47.9%) | 215 (46.4%) | 103 (51.8%) | 108 (46.2%) | 171 (48.7%) |
| Renal failure | 977 (25.4%) | 493 (30.5%) | 319 (25.6%) | 109 (23.5%) | 60 (30.2%) | 65 (27.8%) | 85 (24.2%) |
| Congestive heart failure | 1,270 (33.0%) | 555 (34.4%) | 416 (33.4%) | 149 (32.2%) | 76 (38.2%) | 74 (31.6%) | 117 (33.3%) |
| Chronic pulmonary disease | 712 (18.5%) | 357 (22.1%) | 251 (20.1%) | 84 (18.1%) | 44 (22.1%) | 49 (20.9%) | 74 (21.1%) |
| Obesity | 613 (15.9%) | 213 (13.2%) | 214 (17.2%) | 77 (16.6%) | 44 (22.1%) | 41 (17.5%) | 52 (14.8%) |
| Patients transferred between hospitals | 1,566 (40.7%) | 300 (18.6%) | 498 (39.9%) | 191 (41.3%) | 72 (36.2%) | 97 (41.5%) | 138 (39.3%) |
| Length of hospital stay (days) |  |  |  |  |  |  |  |
| Mean (SD) | 39.4 (34.5) | 21.4 (17.8) | 36.7 (29.7) | 36.7 (30.4) | 36.5 (33.9) | 32.0 (25.2) | 39.9 (28.8) |
| Median (IQR) | 29.0 (16.0, 52.0) | 17.0 (11.0, 27.0) | 28.0 (17.0, 46.5) | 28.0 (16.0, 46.0) | 26.0 (16.0, 46.5) | 26.0 (15.0, 39.0) | 31.0 (20.0, 52.0) |
| Ventilation (days) |  |  |  |  |  |  |  |
| Mean (SD) | 18.6 (17.2) | 4.0 (4.8) | 20.7 (17.3) | 20.0 (16.3) | 21.2 (21.9) | 19.3 (16.1) | 22.3 (16.3) |
| Median (IQR) | 13.7 (6.9, 25.3) | 2.5 (1.1, 5.2) | 16.3 (9.6, 27.6) | 15.0 (8.6, 28.2) | 15.9 (9.9, 25.8) | 15.5 (9.0, 25.2) | 18.1 (11.0, 29.6) |
| Tracheostomy | 1,432 (37.2%) |  | 462 (37.0%) | 165 (35.6%) | 77 (38.7%) | 87 (37.2%) | 133 (37.9%) |
| ECMO | 375 (9.7%) |  | 186 (14.9%) | 68 (14.7%) | 23 (11.6%) | 33 (14.1%) | 62 (17.7%) |
| Dialysis | 1,203 (31.2%) | 75 (4.6%) | 439 (35.2%) | 158 (34.1%) | 70 (35.2%) | 84 (35.9%) | 127 (36.2%) |
| In-hospital mortality | 2,003 (52.0%) | 624 (38.7%) | 818 (65.6%) | 270 (58.3%) | 120 (60.3%) | 165 (70.5%) | 263 (74.9%) |

The duration of MV was clearly dependent on its modality (Table 2). The median duration of MV was 2.5 days in those receiving NIV only but reached 14 days in those who were intubated directly. Of note, patients who were switched from initial NIV to invasive MV following NIV failure spent the longest periods on MV (median 16 days). This trend was also true for the application of ECMO, which was reported in 15% of NIV-F patients, compared to 10% in patients who were intubated without having initially received NIV. Importantly, the proportion of patients with late NIV-F (after 5 days or more of NIV followed by intubation) substantially increased during the second wave, as displayed in Figure 2.

**Figure 2.**
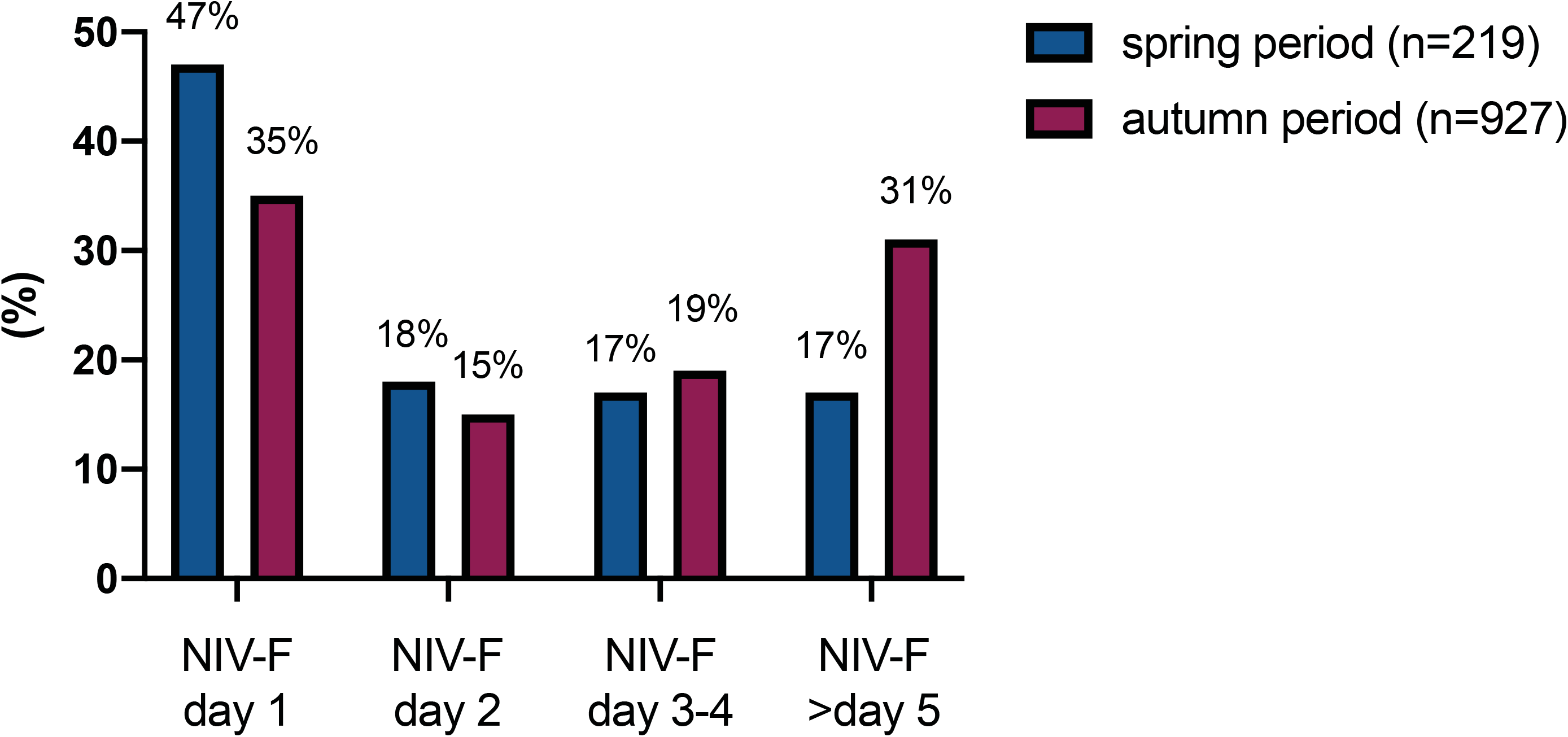
Timely distribution of NIV failure (NIV-F) by day, comparing spring and autumn period.

## Discussion

The current analysis of 7,490 patients represents the largest case series of COVID-19 patients requiring NIV or invasive MV and shows significant differences between the spring and autumn periods 2020 with regard to the modality of MV. The main findings are as follows: Firstly, there was a significant increase in the utilization of NIV in Germany during the second period. Accordingly, the proportion of patients with acute respiratory failure who were directly intubated decreased from 74% to 39%. This was associated with a reduced overall duration of MV, and length of hospital stay. Secondly, the NIV-F rate was still high, even though there was a trend for a lower NIV-F rate during the second period (42%) compared to the first period (49%). Thirdly, the overall mortality rate in patients requiring MV remains high at 53%. Fourthly, NIV-F was associated with an increased ECMO utilization, increased overall duration of MV and increased mortality, and this was particularly true for late NIV-F occurring 5 days or later following NIV initiation.

Several clinical considerations can be derived from the current findings. Most importantly, the present analysis shows that NIV has been clearly established in the treatment of severe respiratory failure attributable to COVID-19 in a real-life setting without resource limitations. Thereby, the overall duration of MV and hospital stay could be shortened. The decreasing NIV-F rate also suggests a learning curve that has occurred over the course of the last year, but may also be related to treatment successes outside MV, such as corticosteroids (22, 23).

The present analysis, however, also demonstrates that clinicians should apply NIV cautiously as NIV-F continues to occur frequently, which is associated with increased mortality. Therefore, NIV certainly cannot replace invasive MV in the majority of MV patients. The rather short median duration of NIV of 2.5 days in those patients successfully treated by NIV suggests that early improvements in respiratory function following NIV identify those patients who have been successfully treated and do not need intubation. In contrast, a longer duration of NIV, particularly exceeding 3–5 days, increases the likelihood of NIV-F, which is associated with an increased mortality.

There are many other reports in the literature also showing the potential of NIV in the treatment of COVID-19-associated respiratory failure (24-26), and this might also have encouraged clinicians to more frequently and extensively apply NIV in this setting. In these reports, NIV was reasonably used outside the ICU, in part aimed at overcoming the shortage of ICU capacities (26). Another rationale to use NIV as long as possible, also in the ICU setting, is aimed at avoiding intubation and intubation-related complications, most importantly lung injury related to invasive MV and infectious complications(8).

In this context, however, clinicians are likely to be less aware of a phenomenon related to maintained spontaneous breathing, which is labeled as patient self-inflicted lung injury (P-SILI) (27). In short, initial lung injury related to COVID-19 is perpetually maintained and even aggravated as a consequence of a vicious circle that includes the sequence of capillary leakage, pulmonary edema, impaired gas exchange and respiratory mechanics, subsequent increase in respiratory drive followed by increased pleural pressure swings, which eventually lead to capillary leakage again if lowering of pleural pressure exceeds the intravascular pressure decrease (28, 29). Even though the current data does not provide evidence for P-SILI in those patients having failed NIV, this phenomenon might, nevertheless, explain why outcome is severely reduced in patients spending longer durations on NIV, which eventually fails.

## Limitations

There are some important limitations of the present paper, which need to be addressed in the context of data interpretation. First, all data refer to the coding of diseases (ICD) and procedures (OPS) in the context of remuneration. Thus, patients were not studied directly. Therefore, several important data are missing, i.e. disease severity related to the PaO_2_/FiO_2_ ratio, intubation criteria, ventilator settings/equipment and oxygen flow rates including information on the response to treatment, “do-not-intubate” orders, details on medication, and the hospital setting (ICU, intermediate care, COVID-19 wards).

Second, the analysis includes only data from one health care insurance company. However, this is the largest insurance and accounts for about 1/3 of the total population, providing a large representative sample for the German population. Third, NIV as defined for the reimbursement system in Germany excludes high-flow oxygen treatment (HFOT) and continuous positive airway pressure (CPAP) and can only be coded if the level of pressure support exceeds 5 cm H_2_O. In addition, the German guidelines have recommended using HFOT as first escalation step when oxygen treatment is insufficient, while CPAP and NIV form the following escalation steps. Thus, NIV in the present analysis represents a rather selected group of patients, and this group may not be compared to studies from other countries without considering this aspect. Finally, patients who have been hospitalized primarily for COVID-19 and those for any other reason in whom COVID-19 could be documented following screening could not be differentiated.

## Conclusions

The utilization of NIV rapidly increased during the autumn period compared to the spring period 2020 of the COVID-19 pandemic in Germany. This was associated with an overall reduced duration of MV, and length of hospital stay. Despite of this, overall mortality of patients receiving MV due to COVID-19-associated respiratory failure remained high at 53%. Patients successfully treated with NIV had lower mortality rates than those who were intubated directly, but those failing NIV had a higher mortality rate, respectively, and this became even more predominant in late NIV failure. Thus, the current study shows the increasing role of NIV in the context of ICU medicine related to COVID-19 and, more specifically, clearly emphasizes its benefits and risks, both impacting on mortality. Given these important findings, there is a need for prospective randomized controlled trials that focus on the most reasonable indications for initiation of NIV as well as timely subsequent intubation in case of NIV failure in COVID-19 patients.

## Data Availability

Data can be viewed upon request.

## References

1. Karagiannidis C, Mostert C, Hentschker C, Voshaar T, Malzahn J, Schillinger G, Klauber J, Janssens U, Marx G, Weber-Carstens S, Kluge S, Pfeifer M, Grabenhenrich L, Welte T, Busse R. Case characteristics, resource use, and outcomes of 10 021 patients with COVID-19 admitted to 920 German hospitals: an observational study. Lancet Respir Med 2020; 8: 853–862.

2. Karagiannidis C, Windisch W, McAuley DF, Welte T, Busse R. Major differences in ICU admissions during the first and second COVID-19 wave in Germany. Lancet Respir Med 2021.

3. Cummings MJ, Baldwin MR, Abrams D, Jacobson SD, Meyer BJ, Balough EM, Aaron JG, Claassen J, Rabbani LE, Hastie J, Hochman BR, Salazar-Schicchi J, Yip NH, Brodie D, O’Donnell MR. Epidemiology, clinical course, and outcomes of critically ill adults with COVID-19 in New York City: a prospective cohort study. Lancet 2020; 395: 1763–1770.

4. Grasselli G, Zangrillo A, Zanella A, Antonelli M, Cabrini L, Castelli A, Cereda D, Coluccello A, Foti G, Fumagalli R, Iotti G, Latronico N, Lorini L, Merler S, Natalini G, Piatti A, Ranieri MV, Scandroglio AM, Storti E, Cecconi M, Pesenti A, Network C-LI, Nailescu A, Corona A, Zangrillo A, Protti A, Albertin A, Forastieri Molinari A, Lombardo A, Pezzi A, Benini A, Scandroglio AM, Malara A, Castelli A, Coluccello A, Micucci A, Pesenti A, Sala A, Alborghetti A, Antonini B, Capra C, Troiano C, Roscitano C, Radrizzani D, Chiumello D, Coppini D, Guzzon D, Costantini E, Malpetti E, Zoia E, Catena E, Agosteo E, Barbara E, Beretta E, Boselli E, Storti E, Harizay F, Della Mura F, Lorini FL, Donato Sigurta F, Marino F, Mojoli F, Rasulo F, Grasselli G, Casella G, De Filippi G, Castelli G, Aldegheri G, Gallioli G, Lotti G, Albano G, Landoni G, Marino G, Vitale G, Battista Perego G, Evasi G, Citerio G, Foti G, Natalini G, Merli G, Sforzini I, Bianciardi L, Carnevale L, Grazioli L, Cabrini L, Guatteri L, Salvi L, Dei Poli M, Galletti M, Gemma M, Ranucci M, Riccio M, Borelli M, Zambon M, Subert M, Cecconi M, Mazzoni MG, Raimondi M, Panigada M, Belliato M, Bronzini N, Latronico N, Petrucci N, Belgiorno N, Tagliabue P, Cortellazzi P, Gnesin P, Grosso P, Gritti P, Perazzo P, Severgnini P, Ruggeri P, Sebastiano P, Covello RD, Fernandez-Olmos R, Fumagalli R, Keim R, Rona R, Valsecchi R, Cattaneo S, Colombo S, Cirri S, Bonazzi S, Greco S, Muttini S, Langer T, Alaimo V, Viola U. Baseline Characteristics and Outcomes of 1591 Patients Infected With SARS-CoV-2 Admitted to ICUs of the Lombardy Region, Italy. JAMA 2020: 1574–1581.

5. Wang Y, Lu X, Li Y, Chen H, Chen T, Su N, Huang F, Zhou J, Zhang B, Yan F, Wang J. Clinical Course and Outcomes of 344 Intensive Care Patients with COVID-19. Am J Respir Crit Care Med 2020; 201: 1430–1434.

6. Sivaloganathan AA, Nasim-Mohi M, Brown MM, Abdul N, Jackson A, Fletcher SV, Gupta S, Grocott MPW, Dushianthan A, University Hospital Southampton Critical C, Respiratory Medicine T, the Ri, Team UHSCCC, Team UHSRC, Investigators R. Noninvasive ventilation for COVID-19-associated acute hypoxaemic respiratory failure: experience from a single centre. Br J Anaesth 2020; 125: e368–e371.

7. Docherty AB, Harrison EM, Green CA, Hardwick HE, Pius R, Norman L, Holden KA, Read JM, Dondelinger F, Carson G, Merson L, Lee J, Plotkin D, Sigfrid L, Halpin S, Jackson C, Gamble C, Horby PW, Nguyen-Van-Tam Js, Ho A, Russell CD, Dunning J, Openshaw PJ, Baillie JK, Semple MG, investigators IC. Features of 20 133 UK patients in hospital with covid-19 using the ISARIC WHO Clinical Characterisation Protocol: prospective observational cohort study. BMJ 2020; 369: m1985.

8. Girou E, Schortgen F, Delclaux C, Brun-Buisson C, Blot F, Lefort Y, Lemaire F, Brochard L. Association of noninvasive ventilation with nosocomial infections and survival in critically ill patients. JAMA 2000; 284: 2361–2367.

9. Raoof S, Nava S, Carpati C, Hill NS. High-Flow, Noninvasive Ventilation and Awake (Nonintubation) Proning in Patients With Coronavirus Disease 2019 With Respiratory Failure. Chest 2020; 158: 1992–2002.

10. Pasin L, Sella N, Correale C, Boscolo A, Rosi P, Saia M, Mantoan D, Navalesi P. Regional COVID-19 Network for Coordination of SARS-CoV-2 outbreak in Veneto, Italy. J Cardiothorac Vasc Anesth 2020; 34: 2341–2345.

11. Kluge S, Janssens U, Spinner CD, Pfeifer M, Marx G, Karagiannidis C, Guideline g. Clinical Practice Guideline: Recommendations on Inpatient Treatment of Patients with COVID-19. Dtsch Arztebl Int 2021; 118.

12. Pfeifer M, Ewig S, Voshaar T, Randerath WJ, Bauer T, Geiseler J, Dellweg D, Westhoff M, Windisch W, Schonhofer B, Kluge S, Lepper PM. Position Paper for the State-of-the-Art Application of Respiratory Support in Patients with COVID-19. Respiration 2020; 99: 521–542.

13. Alhazzani W, Evans L, Alshamsi F, Moller MH, Ostermann M, Prescott HC, Arabi YM, Loeb M, Ng Gong M, Fan E, Oczkowski S, Levy MM, Derde L, Dzierba A, Du B, Machado F, Wunsch H, Crowther M, Cecconi M, Koh Y, Burry L, Chertow DS, Szczeklik W, Belley-Cote E, Greco M, Bala M, Zarychanski R, Kesecioglu J, McGeer A, Mermel L, Mammen MJ, Nainan Myatra S, Arrington A, Kleinpell R, Citerio G, Lewis K, Bridges E, Memish ZA, Hammond N, Hayden FG, Alshahrani M, Al Duhailib Z, Martin GS, Kaplan LJ, Coopersmith CM, Antonelli M, Rhodes A. Surviving Sepsis Campaign Guidelines on the Management of Adults With Coronavirus Disease 2019 (COVID-19) in the ICU: First Update. Crit Care Med 2021; 49: e219–e234.

14. Vianello A, Arcaro G, Molena B, Turato C, Sukthi A, Guarnieri G, Lugato F, Senna G, Navalesi P. High-flow nasal cannula oxygen therapy to treat patients with hypoxemic acute respiratory failure consequent to SARS-CoV-2 infection. Thorax 2020; 75: 998–1000.

15. Fan E, Beitler JR, Brochard L, Calfee CS, Ferguson ND, Slutsky AS, Brodie D. COVID-19-associated acute respiratory distress syndrome: is a different approach to management warranted? Lancet Respir Med 2020; 8: 816–821.

16. Alqahtani JS, Mendes RG, Aldhahir A, Rowley D, AlAhmari MD, Ntoumenopoulos G, Alghamdi SM, Sreedharan JK, Aldabayan YS, Oyelade T, Alrajeh A, Olivieri C, AlQuaimi M, Sullivan J, Almeshari MA, Esquinas A. Global Current Practices of Ventilatory Support Management in COVID-19 Patients: An International Survey. J Multidiscip Healthc 2020; 13: 1635–1648.

17. Network C-IGobotR, the C-ICUI. Clinical characteristics and day-90 outcomes of 4244 critically ill adults with COVID-19: a prospective cohort study. Intensive Care Med 2021; 47: 60–73.

18. Bellani G, Grasselli G, Cecconi M, Antolini L, Borelli M, De Giacomi F, Bosio G, Latronico N, Filippini M, Gemma M, Giannotti C, Antonini B, Petrucci N, Zerbi SM, Maniglia P, Castelli GP, Marino G, Subert M, Citerio G, Radrizzani D, Mediani TS, Lorini FL, Russo FM, Faletti A, Beindorf A, Covello RD, Greco S, Bizzarri MM, Ristagno G, Mojoli F, Pradella A, Severgnini P, Da Macalle M, Albertin A, Ranieri VM, Rezoagli E, Vitale G, Magliocca A, Cappelleri G, Docci M, Aliberti S, Serra F, Rossi E, Valsecchi MG, Pesenti A, Foti G, Network C-LI. Noninvasive Ventilatory Support of COVID-19 Patients Outside the Intensive Care Units (WARd-COVID). Ann Am Thorac Soc 2021.

19. Oranger M, Gonzalez-Bermejo J, Dacosta-Noble P, Llontop C, Guerder A, Trosini-Desert V, Faure M, Raux M, Decavele M, Demoule A, Morelot-Panzini C, Similowski T. Continuous positive airway pressure to avoid intubation in SARS-CoV-2 pneumonia: a two-period retrospective case-control study. Eur Respir J 2020; 56.

20. Aliberti S, Radovanovic D, Billi F, Sotgiu G, Costanzo M, Pilocane T, Saderi L, Gramegna A, Rovellini A, Perotto L, Monzani V, Santus P, Blasi F. Helmet CPAP treatment in patients with COVID-19 pneumonia: a multicentre cohort study. Eur Respir J 2020; 56.

21. Tuffet S, Mekontso Dessap A, Carteaux G. Noninvasive Ventilation for De Novo Respiratory Failure: Impact of Ventilator Setting Adjustments. Am J Respir Crit Care Med 2020; 202: 769–770.

22. Group RC, Horby P, Lim WS, Emberson JR, Mafham M, Bell JL, Linsell L, Staplin N, Brightling C, Ustianowski A, Elmahi E, Prudon B, Green C, Felton T, Chadwick D, Rege K, Fegan C, Chappell LC, Faust SN, Jaki T, Jeffery K, Montgomery A, Rowan K, Juszczak E, Baillie JK, Haynes R, Landray MJ. Dexamethasone in Hospitalized Patients with Covid-19 - Preliminary Report. N Engl J Med 2020.

23. Angus DC, Derde L, Al-Beidh F, Annane D, Arabi Y, Beane A, van Bentum-Puijk W, Berry L, Bhimani Z, Bonten M, Bradbury C, Brunkhorst F, Buxton M, Buzgau A, Cheng AC, de Jong M, Detry M, Estcourt L, Fitzgerald M, Goossens H, Green C, Haniffa R, Higgins AM, Horvat C, Hullegie SJ, Kruger P, Lamontagne F, Lawler PR, Linstrum K, Litton E, Lorenzi E, Marshall J, McAuley D, McGlothin A, McGuinness S, McVerry B, Montgomery S, Mouncey P, Murthy S, Nichol A, Parke R, Parker J, Rowan K, Sanil A, Santos M, Saunders C, Seymour C, Turner A, van de Veerdonk F, Venkatesh B, Zarychanski R, Berry S, Lewis RJ, McArthur C, Webb SA, Gordon AC, Writing Committee for the R-CAPI, Al-Beidh F, Angus D, Annane D, Arabi Y, van Bentum-Puijk W, Berry S, Beane A, Bhimani Z, Bonten M, Bradbury C, Brunkhorst F, Buxton M, Cheng A, De Jong M, Derde L, Estcourt L, Goossens H, Gordon A, Green C, Haniffa R, Lamontagne F, Lawler P, Litton E, Marshall J, McArthur C, McAuley D, McGuinness S, McVerry B, Montgomery S, Mouncey P, Murthy S, Nichol A, Parke R, Rowan K, Seymour C, Turner A, van de Veerdonk F, Webb S, Zarychanski R, Campbell L, Forbes A, Gattas D, Heritier S, Higgins L, Kruger P, Peake S, Presneill J, Seppelt I, Trapani T, Young P, Bagshaw S, Daneman N, Ferguson N, Misak C, Santos M, Hullegie S, Pletz M, Rohde G, Rowan K, Alexander B, Basile K, Girard T, Horvat C, Huang D, Linstrum K, Vates J, Beasley R, Fowler R, McGloughlin S, Morpeth S, Paterson D, Venkatesh B, Uyeki T, Baillie K, Duffy E, Fowler R, Hills T, Orr K, Patanwala A, Tong S, Netea M, Bihari S, Carrier M, Fergusson D, Goligher E, Haidar G, Hunt B, Kumar A, Laffan M, Lawless P, Lother S, McCallum P, Middeldopr S, McQuilten Z, Neal M, Pasi J, Schutgens R, Stanworth S, Turgeon A, Weissman A, Adhikari N, Anstey M, Brant E, de Man A, Lamonagne F, Masse MH, Udy A, Arnold D, Begin P, Charlewood R, Chasse M, Coyne M, Cooper J, Daly J, Gosbell I, Harvala-Simmonds H, Hills T, MacLennan S, Menon D, McDyer J, Pridee N, Roberts D, Shankar-Hari M, Thomas H, Tinmouth A, Triulzi D, Walsh T, Wood E, Calfee C, O’Kane C, Shyamsundar M, Sinha P, Thompson T, Young I, Bihari S, Hodgson C, Laffey J, McAuley D, Orford N, Neto A, Detry M, Fitzgerald M, Lewis R, McGlothlin A, Sanil A, Saunders C, Berry L, Lorenzi E, Miller E, Singh V, Zammit C, van Bentum Puijk W, Bouwman W, Mangindaan Y, Parker L, Peters S, Rietveld I, Raymakers K, Ganpat R, Brillinger N, Markgraf R, Ainscough K, Brickell K, Anjum A, Lane JB, Richards-Belle A, Saull M, Wiley D, Bion J, Connor J, Gates S, Manax V, van der Poll T, Reynolds J, van Beurden M, Effelaar E, Schotsman J, Boyd C, Harland C, Shearer A, Wren J, Clermont G, Garrard W, Kalchthaler K, King A, Ricketts D, Malakoutis S, Marroquin O, Music E, Quinn K, Cate H, Pearson K, Collins J, Hanson J, Williams P, Jackson S, Asghar A, Dyas S, Sutu M, Murphy S, Williamson D, Mguni N, Potter A, Porter D, Goodwin J, Rook C, Harrison S, Williams H, Campbell H, Lomme K, Williamson J, Sheffield J, van’t Hoff W, McCracken P, Young M, Board J, Mart E, Knott C, Smith J, Boschert C, Affleck J, Ramanan M, D’Souza R, Pateman K, Shakih A, Cheung W, Kol M, Wong H, Shah A, Wagh A, Simpson J, Duke G, Chan P, Cartner B, Hunter S, Laver R, Shrestha T, Regli A, Pellicano A, McCullough J, Tallott M, Kumar N, Panwar R, Brinkerhoff G, Koppen C, Cazzola F, Brain M, Mineall S, Fischer R, Biradar V, Soar N, White H, Estensen K, Morrison L, Smith J, Cooper M, Health M, Shehabi Y, Al-Bassam W, Hulley A, Whitehead C, Lowrey J, Gresha R, Walsham J, Meyer J, Harward M, Venz E, Williams P, Kurenda C, Smith K, Smith M, Garcia R, Barge D, Byrne D, Byrne K, Driscoll A, Fortune L, Janin P, Yarad E, Hammond N, Bass F, Ashelford A, Waterson S, Wedd S, McNamara R, Buhr H, Coles J, Schweikert S, Wibrow B, Rauniyar R, Myers E, Fysh E, Dawda A, Mevavala B, Litton E, Ferrier J, Nair P, Buscher H, Reynolds C, Santamaria J, Barbazza L, Homes J, Smith R, Murray L, Brailsford J, Forbes L, Maguire T, Mariappa V, Smith J, Simpson S, Maiden M, Bone A, Horton M, Salerno T, Sterba M, Geng W, Depuydt P, De Waele J, De Bus L, Fierens J, Bracke S, Reeve B, Dechert W, Chasse M, Carrier FM, Boumahni D, Benettaib F, Ghamraoui A, Bellemare D, Cloutier E, Francoeur C, Lamontagne F, D’Aragon F, Carbonneau E, Leblond J, Vazquez-Grande G, Marten N, Wilson M, Albert M, Serri K, Cavayas A, Duplaix M, Williams V, Rochwerg B, Karachi T, Oczkowski S, Centofanti J, Millen T, Duan E, Tsang J, Patterson L, English S, Watpool I, Porteous R, Miezitis S, McIntyre L, Brochard L, Burns K, Sandhu G, Khalid I, Binnie A, Powell E, McMillan A, Luk T, Aref N, Andric Z, Cviljevic S, Dimoti R, Zapalac M, Mirkovic G, Barsic B, Kutlesa M, Kotarski V, Vujaklija Brajkovic A, Babel J, Sever H, Dragija L, Kusan I, Vaara S, Pettila L, Heinonen J, Kuitunen A, Karlsson S, Vahtera A, Kiiski H, Ristimaki S, Azaiz A, Charron C, Godement M, Geri G, Vieillard-Baron A, Pourcine F, Monchi M, Luis D, Mercier R, Sagnier A, Verrier N, Caplin C, Siami S, Aparicio C, Vautier S, Jeblaoui A, Fartoukh M, Courtin L, Labbe V, Leparco C, Muller G, Nay MA, Kamel T, Benzekri D, Jacquier S, Mercier E, Chartier D, Salmon C, Dequin P, Schneider F, Morel G, L’Hotellier S, Badie J, Berdaguer FD, Malfroy S, Mezher C, Bourgoin C, Megarbane B, Voicu S, Deye N, Malissin I, Sutterlin L, Guitton C, Darreau C, Landais M, Chudeau N, Robert A, Moine P, Heming N, Maxime V, Bossard I, Nicholier TB, Colin G, Zinzoni V, Maquigneau N, Finn A, Kress G, Hoff U, Friedrich Hinrichs C, Nee J, Pletz M, Hagel S, Ankert J, Kolanos S, Bloos F, Petros S, Pasieka B, Kunz K, Appelt P, Schutze B, Kluge S, Nierhaus A, Jarczak D, Roedl K, Weismann D, Frey A, Klinikum Neukolln V, Reill L, Distler M, Maselli A, Belteczki J, Magyar I, Fazekas A, Kovacs S, Szoke V, Szigligeti G, Leszkoven J, Collins D, Breen P, Frohlich S, Whelan R, McNicholas B, Scully M, Casey S, Kernan M, Doran P, O’Dywer M, Smyth M, Hayes L, Hoiting O, Peters M, Rengers E, Evers M, Prinssen A, Bosch Ziekenhuis J, Simons K, Rozendaal W, Polderman F, de Jager P, Moviat M, Paling A, Salet A, Rademaker E, Peters AL, de Jonge E, Wigbers J, Guilder E, Butler M, Cowdrey KA, Newby L, Chen Y, Simmonds C, McConnochie R, Ritzema Carter J, Henderson S, Van Der Heyden K, Mehrtens J, Williams T, Kazemi A, Song R, Lai V, Girijadevi D, Everitt R, Russell R, Hacking D, Buehner U, Williams E, Browne T, Grimwade K, Goodson J, Keet O, Callender O, Martynoga R, Trask K, Butler A, Schischka L, Young C, Lesona E, Olatunji S, Robertson Y, Jose N, Amaro dos Santos Catorze T, de Lima Pereira Tna, Neves Pessoa LM, Castro Ferreira RM, Pereira Sousa Bastos JM, Aysel Florescu S, Stanciu D, Zaharia MF, Kosa AG, Codreanu D, Marabi Y, Al Qasim E, Moneer Hagazy M, Al Swaidan L, Arishi H, Munoz-Bermudez R, Marin-Corral J, Salazar Degracia A, Parrilla Gomez F, Mateo Lopez MI, Rodriguez Fernandez J, Carcel Fernandez S, Carmona Flores R, Leon Lopez R, de la Fuente Martos C, Allan A, Polgarova P, Farahi N, McWilliam S, Hawcutt D, Rad L, O’Malley L, Whitbread J, Kelsall O, Wild L, Thrush J, Wood H, Austin K, Donnelly A, Kelly M, O’Kane S, McClintock D, Warnock M, Johnston P, Gallagher LJ, Mc Goldrick C, Mc Master M, Strzelecka A, Jha R, Kalogirou M, Ellis C, Krishnamurthy V, Deelchand V, Silversides J, McGuigan P, Ward K, O’Neill A, Finn S, Phillips B, Mullan D, Oritz-Ruiz de Gordoa L, Thomas M, Sweet K, Grimmer L, Johnson R, Pinnell J, Robinson M, Gledhill L, Wood T, Morgan M, Cole J, Hill H, Davies M, Antcliffe D, Templeton M, Rojo R, Coghlan P, Smee J, Mackay E, Cort J, Whileman A, Spencer T, Spittle N, Kasipandian V, Patel A, Allibone S, Genetu RM, Ramali M, Ghosh A, Bamford P, London E, Cawley K, Faulkner M, Jeffrey H, Smith T, Brewer C, Gregory J, Limb J, Cowton A, O’Brien J, Nikitas N, Wells C, Lankester L, Pulletz M, Williams P, Birch J, Wiseman S, Horton S, Alegria A, Turki S, Elsefi T, Crisp N, Allen L, McCullagh I, Robinson P, Hays C, Babio-Galan M, Stevenson H, Khare D, Pinder M, Selvamoni S, Gopinath A, Pugh R, Menzies D, Mackay C, Allan E, Davies G, Puxty K, McCue C, Cathcart S, Hickey N, Ireland J, Yusuff H, Isgro G, Brightling C, Bourne M, Craner M, Watters M, Prout R, Davies L, Pegler S, Kyeremeh L, Arbane G, Wilson K, Gomm L, Francia F, Brett S, Sousa Arias S, Elin Hall R, Budd J, Small C, Birch J, Collins E, Henning J, Bonner S, Hugill K, Cirstea E, Wilkinson D, Karlikowski M, Sutherland H, Wilhelmsen E, Woods J, North J, Sundaran D, Hollos L, Coburn S, Walsh J, Turns M, Hopkins P, Smith J, Noble H, Depante MT, Clarey E, Laha S, Verlander M, Williams A, Huckle A, Hall A, Cooke J, Gardiner-Hill C, Maloney C, Qureshi H, Flint N, Nicholson S, Southin S, Nicholson A, Borgatta B, Turner-Bone I, Reddy A, Wilding L, Chamara Warnapura L, Agno Sathianathan R, Golden D, Hart C, Jones J, Bannard-Smith J, Henry J, Birchall K, Pomeroy F, Quayle R, Makowski A, Misztal B, Ahmed I, KyereDiabour T, Naiker K, Stewart R, Mwaura E, Mew L, Wren L, Willams F, Innes R, Doble P, Hutter J, Shovelton C, Plumb B, Szakmany T, Hamlyn V, Hawkins N, Lewis S, Dell A, Gopal S, Ganguly S, Smallwood A, Harris N, Metherell S, Lazaro JM, Newman T, Fletcher S, Nortje J, Fottrell-Gould D, Randell G, Zaman M, Elmahi E, Jones A, Hall K, Mills G, Ryalls K, Bowler H, Sall J, Bourne R, Borrill Z, Duncan T, Lamb T, Shaw J, Fox C, Moreno Cuesta J, Xavier K, Purohit D, Elhassan M, Bakthavatsalam D, Rowland M, Hutton P, Bashyal A, Davidson N, Hird C, Chhablani M, Phalod G, Kirkby A, Archer S, Netherton K, Reschreiter H, Camsooksai J, Patch S, Jenkins S, Pogson D, Rose S, Daly Z, Brimfield L, Claridge H, Parekh D, Bergin C, Bates M, Dasgin J, McGhee C, Sim M, Hay SK, Henderson S, Phull MK, Zaidi A, Pogreban T, Rosaroso LP, Harvey D, Lowe B, Meredith M, Ryan L, Hormis A, Walker R, Collier D, Kimpton S, Oakley S, Rooney K, Rodden N, Hughes E, Thomson N, McGlynn D, Walden A, Jacques N, Coles H, Tilney E, Vowell E, Schuster-Bruce M, Pitts S, Miln R, Purandare L, Vamplew L, Spivey M, Bean S, Burt K, Moore L, Day C, Gibson C, Gordon E, Zitter L, Keenan S, Baker E, Cherian S, Cutler S, Roynon-Reed A, Harrington K, Raithatha A, Bauchmuller K, Ahmad N, Grecu I, Trodd D, Martin J, Wrey Brown C, Arias AM, Craven T, Hope D, Singleton J, Clark S, Rae N, Welters I, Hamilton DO, Williams K, Waugh V, Shaw D, Puthucheary Z, Martin T, Santos F, Uddin R, Somerville A, Tatham KC, Jhanji S, Black E, Dela Rosa A, Howle R, Tully R, Drummond A, Dearden J, Philbin J, Munt S, Vuylsteke A, Chan C, Victor S, Matsa R, Gellamucho M, Creagh-Brown B, Tooley J, Montague L, De Beaux F, Bullman L, Kersiake I, Demetriou C, Mitchard S, Ramos L, White K, Donnison P, Johns M, Casey R, Mattocks L, Salisbury S, Dark P, Claxton A, McLachlan D, Slevin K, Lee S, Hulme J, Joseph S, Kinney F, Senya HJ, Oborska A, Kayani A, Hadebe B, Orath Prabakaran R, Nichols L, Thomas M, Worner R, Faulkner B, Gendall E, Hayes K, Hamilton-Davies C, Chan C, Mfuko C, Abbass H, Mandadapu V, Leaver S, Forton D, Patel K, Paramasivam E, Powell M, Gould R, Wilby E, Howcroft C, Banach D, Fernandez de Pinedo Artaraz Z, Cabreros L, White I, Croft M, Holland N, Pereira R, Zaki A, Johnson D, Jackson M, Garrard H, Juhaz V, Roy A, Rostron A, Woods L, Cornell S, Pillai S, Harford R, Rees T, Ivatt H, Sundara Raman A, Davey M, Lee K, Barber R, Chablani M, Brohi F, Jagannathan V, Clark M, Purvis S, Wetherill B, Dushianthan A, Cusack R, de Courcy-Golder K, Smith S, Jackson S, Attwood B, Parsons P, Page V, Zhao XB, Oza D, Rhodes J, Anderson T, Morris S, Xia Le Tai C, Thomas A, Keen A, Digby S, Cowley N, Wild L, Southern D, Reddy H, Campbell A, Watkins C, Smuts S, Touma O, Barnes N, Alexander P, Felton T, Ferguson S, Sellers K, Bradley-Potts J, Yates D, Birkinshaw I, Kell K, Marshall N, Carr-Knott L. Effect of Hydrocortisone on Mortality and Organ Support in Patients With Severe COVID-19: The REMAP-CAP COVID-19 Corticosteroid Domain Randomized Clinical Trial. JAMA 2020; 324: 1317–1329.

24. Coppo A, Bellani G, Winterton D, Di Pierro M, Soria A, Faverio P, Cairo M, Mori S, Messinesi G, Contro E, Bonfanti P, Benini A, Valsecchi MG, Antolini L, Foti G. Feasibility and physiological effects of prone positioning in non-intubated patients with acute respiratory failure due to COVID-19 (PRON-COVID): a prospective cohort study. Lancet Respir Med 2020; 8: 765–774.

25. Sartini C, Tresoldi M, Scarpellini P, Tettamanti A, Carco F, Landoni G, Zangrillo A. Respiratory Parameters in Patients With COVID-19 After Using Noninvasive Ventilation in the Prone Position Outside the Intensive Care Unit. JAMA 2020; 323: 2338–2340.

26. Franco C, Facciolongo N, Tonelli R, Dongilli R, Vianello A, Pisani L, Scala R, Malerba M, Carlucci A, Negri EA, Spoladore G, Arcaro G, Tillio PA, Lastoria C, Schifino G, Tabbi L, Guidelli L, Guaraldi G, Ranieri VM, Clini E, Nava S. Feasibility and clinical impact of out-of-ICU noninvasive respiratory support in patients with COVID-19-related pneumonia. Eur Respir J 2020; 56.

27. Brochard L, Slutsky A, Pesenti A. Mechanical Ventilation to Minimize Progression of Lung Injury in Acute Respiratory Failure. Am J Respir Crit Care Med 2017; 195: 438–442.

28. Windisch W, Weber-Carstens S, Kluge S, Rossaint R, Welte T, Karagiannidis C. Invasive and Non-Invasive Ventilation in Patients With COVID-19. Dtsch Arztebl Int 2020; 117: 528–533.

29. Tonelli R, Fantini R, Tabbi L, Castaniere I, Pisani L, Pellegrino MR, Della Casa G, D’Amico R, Girardis M, Nava S, Clini EM, Marchioni A. Early Inspiratory Effort Assessment by Esophageal Manometry Predicts Noninvasive Ventilation Outcome in De Novo Respiratory Failure. A Pilot Study. Am J Respir Crit Care Med 2020; 202: 558–567.

